# Behavioral and structural barriers to human post-exposure prophylaxis and other preventive practices during a canine rabies epidemic

**DOI:** 10.1101/2020.01.16.20016394

**Authors:** Ricardo Castillo-Neyra, Alison M. Buttenheim, Joanna Brown, James F. Ferrara, Claudia Arevalo-Nieto, Katty Borrini-Mayorí, Michael Z. Levy, Victor Becerra, Valerie A. Paz-Soldan

**Affiliations:** Department of Biostatistics, Epidemiology & Informatics, Perelman School of Medicine at University of Pennsylvania, Philadelphia, Pennsylvania, United States of America; Zoonotic Disease Research Lab, One Health Unit, School of Public Health and Administration, Universidad Peruana Cayetano Heredia, Lima, Peru; Department of Family and Community Health, University of Pennsylvania School of Nursing, Philadelphia, Pennsylvania, United States of America; School of Veterinary Medicine, University of Pennsylvania, Pennsylvania, United States of America; Microred Mariano Melgar, Ministerio de Salud, Arequipa, Peru; Department of Global Community Health and Behavioral Sciences, Tulane University School of Public Health and Tropical Medicine, New Orleans, Louisiana, United States

**Keywords:** Focus groups, human rabies, post-exposure prophylaxis, qualitative research, rabies outbreak, urban rabies, zoonosis

## Abstract

A canine rabies epidemic started in early 2015 in Arequipa, Peru; the rabies virus continues to circulate in the dog population. Some city residents who suffer dog bites do not seek care or do not complete indicated post-exposure prophylaxis (PEP) regimens, increasing the risk of human rabies. The objectives of our study were to qualitatively assess knowledge about rabies, and preventive practices, such as PEP vaccination, following a dog bite. We conducted eight focus group discussions in peri-urban and urban communities with 70 total participants. We observed low awareness of rabies severity and fatality. Participants, especially those in per-urban communities, recounted applying herbs or the hair of the dog that bit them to wounds rather than seeking appropriate care. Misconceptions about rabies vaccines and mistreatment at health centers also commonly prevents initiating or completing PEP vaccination. We identify important behavioral and structural barriers and knowledge gaps that limit evidence-based preventive strategies against rabies and may threaten successful prevention of dog-mediated human rabies in this setting.

## Introduction

The city of Arequipa, Peru has been in the midst of a canine rabies epidemic for the past five years (1,2). Fortunately, no human cases have yet been detected. However, continued transmission of the rabies virus in the dog population puts the one million inhabitants of the city at risk of infection. The Peruvian Ministry of Health (MOH) has taken steps since the beginning of the outbreak to prevent human transmission. They have conducted mass dog vaccination campaigns, coordinated health promotion campaigns, and provided, at no cost, post-exposure prophylaxis (PEP) vaccination based on cell-derived vaccines (3) to people exposed or potentially exposed to the rabies virus. However, health posts report that some exposed residents do not seek professional care for bites or follow the PEP vaccination regimen to completion, thereby increasing the risk of human rabies in Arequipa. In one of the 14 districts in the city, up to 14% of residents reported being bitten by a dog of unknown vaccination status during the first year of the outbreak; of these, only 22% sought medical care (unpublished). We do not know how many residents bitten by a dog start the PEP regimen but do not complete it; healthcare providers and authorities from the region contacted us concerned over incomplete PEP regimens and wanting to understand more about possible reasons for not adhering to the five shot sequence.

Rabies has the highest case-fatality ratio of any infectious disease (4); once the disease manifests clinically, the outcome is almost invariably fatal (5,6). However, human rabies is preventable through a well-characterized protocol, including thorough washing of all wounds with soap and copious amounts of water, prompt initiation and completion of PEP, and administration of rabies immunoglobulin if indicated and available (7-9). The PEP regimen varies based on the type of vaccine; in Peru, the vaccine regimen during the development of this study consisted of 5 intramuscular shots given at days 0, 3, 7, 14, and 28 and was provided for free by the Ministry of Health (10).

Despite high fatality and effective prophylaxis, people do not always seek medical care following a bite. Those that do initiate PEP swiftly often do not complete it (11,12). Various factors contribute to limited or delayed treatment seeking, including the lack of perceived need for immediate treatment after exposure (13), preference for traditional medicine, giving priority to earning a livelihood over seeking care (14), and increasingly, vaccine hesitancy (15,16). Some report that rabies vaccine injections are painful and difficult to tolerate (17); parents report desperation when they see their kids in pain because of a vaccine (15). In some settings (although not in Peru), rabies vaccines are expensive, creating a social justice issue for bite victims and their communities who are often least able to afford care and most vulnerable to rabies mortality (18). Unfortunately, despite aggressive and successful dog vaccination programs (19) and widely-available human post-exposure prophylaxis, 59,000 humans die of rabies every year (20).

The plentiful literature on rabies transmission and PEP compliance highlights gaps in rabies knowledge and in seeking medical treatment, including PEP, especially when urgently indicated (1,2,6-8,20-23). Most studies show that respondents recognize the dog as a primary reservoir for rabies transmission and that the majority of respondents agree that rabies can be transmitted by a dog bite, but fewer individuals recognize that transmission is through the dog’s saliva (1,2,4,5,7,8,19,20). Some studies have identified poorer knowledge of rabies in urban vs. non-urban populations (5,19,20). Individual factors associated with poorer rabies knowledge include being single, having more children, lower educational attainment, having a sedentary lifestyle, and being younger (5,19,20). Variable knowledge of the importance of using soap and water for bite wounds has been reported. Around half the population surveyed in Western India was aware of the importance of washing a wound, compared to a very small percentage of those surveyed in Tanzania (6,7,19,20). A majority of those surveyed who used alternative medicine, such as turmeric powder in India, had a delayed hospital presentation (> 24 hours) (14,24). That said, increasing knowledge about rabies and its prevention is likely necessary but not sufficient for behavior change (1,2,6-8,20).

In general, rabies mortality has been associated with inadequate wound management, delayed arrival to the healthcare services, and misconceptions about animal bites (14), and these associations are more prevalent in non-urban areas compared to urban ones (13,14,17,24-28). The likelihood of seeking care for a bite or receiving a PEP recommendation depended on factors including: which animal (dog vs. cat) bit the person; whether the biting animal was owned (vs. unowned), whether the person knew the vaccination status of the biting animal, and the rabies and PEP knowledge of the medical provider (1,2,4). If the bite was from a dog, especially if unowned or unvaccinated (4), PEP was more likely to be requested and/or recommended. In survey-based studies, while the majority of respondents knew the mortality risk of rabies (1,2,7,19-21), only a minority of those who identified as bite victims actually sought out a full rabies vaccination series (2,21). Significant individual-level characteristics associated with lack of PEP knowledge and practices included no formal education, owning only one dog, and living far from a vaccination site (e.g. health facility) (6,7).

In a city with a re-emergent epidemic of canine rabies where low uptake and low completion of PEP vaccination is reported by health practitioners, the objectives of this study were to: 1) qualitatively assess people’s knowledge and practices following any dog bite, 2) characterize differences in prevention perceptions and strategies by levels of urbanization, and 3) examine community attitudes towards PEP vaccination, and 4) potential behavioral and structural barriers to initiating and completing PEP. Focus groups were conducted in urban and peri-urban communities, and during the analysis, the results were stratified by these two groups to examine potential differences in their prevention perceptions and strategies. Findings from this study can improve the framing and content of health communication campaigns to increase appropriate health-care seeking practices, with a focus on seeking and completing PEP.

## 2. Materials and Methods

### 2.1. Ethics statement

This study was approved by the Institutional Review Boards from Universidad Peruana Cayetano Heredia (approval identification number: 65369), Tulane University (#14-606720), and University of Pennsylvania (#823736).

### 2.2. Study settings and study population

This study was conducted in the Mariano Melgar (MM) district of Arequipa. The city of Arequipa—the second largest in Peru—has a population of 969,000 and is situated ∼2300 meters above sea level (29). MM has 14,500 households and like many districts in Arequipa, has experienced heterogeneous growth and urban development, related to significant unplanned immigration to this city over several decades. Half of the rabid dogs detected in Arequipa have been detected in MM.

Participants in our study came from both urban and peri-urban areas of MM. Urban neighborhoods were founded several decades (in some cases, centuries) ago, and tend to have wealthier and more highly-educated residents compared to those in the peri-urban areas. Homes tend to be more secure due to better construction materials in the urban areas compared to those in the peri-urban areas. Peri-urban neighborhoods, founded just 10-50 years ago during a time of mass rural to urban migration in Peru, tend to have housing made from cheap and often temporary materials, fewer community resources, and more security problems (2).

### 2.3. Study Design

Data for this qualitative study were obtained through focus group discussions (FGD). The research team developed and applied a focus group guide aimed at examining the following topics: 1) knowledge of rabies virus transmission in dogs and humans, 2) knowledge of signs of canine rabies, 3) knowledge of and practices associated to management of dog bite wounds, and 4) knowledge about and awareness of post-exposure prophylaxis (PEP), and the importance of its completion. Additional topics in the interview guide included dog vaccination and dog ownership; these were analyzed separately and results described elsewhere (2).

Three Peruvian facilitators conducted the FGD: VPS, a social scientist, PhD in public health and experienced focus group facilitator; JBr, BA in psychology and research assistant; and RCN, infectious disease scientist, veterinarian and PhD in epidemiology who leads the study in Arequipa. Another team member from Arequipa took extensive notes. All FGD were carried out in January 2016. After each focus group, a 15-minute conversation was held with the participants to answer questions and eliminate misconceptions about rabies. If any of the FGD participants reported taking actions that would warrant medical care we would refer to the health center for same-day medical advice.

### 2.4. Sampling strategy

Participants for the eight FGD were recruited from urban and peri-urban neighborhoods of MM through purposive sampling. Participants in households close to health facilities and the district municipal office building are often more exposed to rabies vaccination campaign messages or general health promotion information. Therefore, participants were recruited from households at least 6 blocks away from these sites by pre-selecting blocks on a map.

### 2.5. Recruitment approach

Recruitment was done by visiting every third house door-to-door in the pre-selected blocks the day before the FGD to recruit 2 or 3 participants per block willing to participate and who met the inclusion criteria. The total number of desired participants per FGD (8 to 12) was reached applying the same procedure in adjacent blocks, although given that the turnout might be low, up to 15 participants were invited. While picking up participants by hired van 15 minutes prior to the FGD, the field team reviewed the informed consent forms with participants to ensure their interest in participating. Participants were picked up from their households with hired transportation 10 to 15 minutes before the FGD and were compensated for their transport home. We recruited a diverse sample (by gender, adult age group, and dog ownership status), ensuring diversity of perceptions and experiences. Prior to starting the FGD, the research team once again reviewed the informed consent with the participants and obtained written consent from each to participate for audio recording.

### 2.6. Data management and analysis

Digital audio recordings and written notes from the eight FGDs were transcribed verbatim. An inductive coding approach was applied to identify topics that emerged from the data (30). Codes were generated and organized based on emergent and *a priori* themes of interest related to behavioral and structural barriers to PEP and other prevention strategies. Coding was done by three authors (JBr, CAN, KBM) using ATLAS.ti (31) software. Inconsistent coding was flagged, discussed, and resolved, returning to transcripts when necessary. The analysis of the principal topics of interest was stratified by urban and peri-urban communities, but only mentioned in text when we found differences. All de-identified data are available at https://github.com/chirimacha/Rabies_PEP_Arequipa_qualitative.

## 3. Results

The number of participants per focus group ranged from 7 to 11, with a total N of 70. Participants were predominantly female (77%) and dog owners (91%). Most participants from the urban communities were born in the city of Arequipa (75%) versus participants from peri-urban who were mostly born in the countryside of Arequipa or in other Peruvian states (63%). Half of all participants were housewives or reported working at home, 36% reported working out of the house in jobs spanning from law firms to farms, 8% were students, and 6% were unemployed or did not disclose their occupation.

### 3.1. Knowledge of rabies virus transmission in dogs and humans

When asked how dogs become infected with rabies, “dog bites” were identified by most in both the urban and peri-urban areas. However, participants also reported that rabies could be lingering in the environment, and that certain conditions—specifically heat, overexposure to sun, and lack of water for the dog—could lead to dogs acquiring rabies:

> *“It is my understanding that rabies occurs because the dog is too much in the sun, and that happens more when the dog is on the street, less when it’s at home*.*” (Female, peri-urban area)*

Participants in both areas also mentioned that flies or rats can infect dogs, possibly by direct contact with an open wound. Parasites, poor diet, the stress and frustration of being tied up, and isolation or lack of contact with people were also cited as risk factors for rabies:

> *“The dog wants to talk but it can’t, so there’s impotence; I imagine it’s stressed. Poor dog. And that’s why they produce rabies*.*” (Female, urban area)*

Although most participants correctly recognized bites from dogs as the main source of transmission to humans, none mentioned dog scratches as possible transmission mechanisms. One urban resident specifically expressed doubts about saliva playing a role in transmission.

FGD participants also differentiated risk of rabies transmission by the dog’s ownership status (owned vs. stray). There was a common perception that stray dogs are at particular risk of carrying rabies because they are not vaccinated, enter easily into fights, are dirty, and often carry other diseases such as scabies. This led to speculation about whether it is necessary to vaccinate dogs that are not let out of the home:

> *“… if my dog is in the house all day, how will it have rabies if it doesn’t get together with stray dogs? My dog is not from the street, it’s only in the house, it’s domestic. So… maybe I won’t give [worrying about whether her dog may get rabies] much importance*.*”*

(Female, peri-urban area).

FGD participants from both areas suggested there should be more information about human rabies on television, including people’s testimonies, to improve dissemination about disease prevention.

### 3.2. Knowledge of signs and symptoms of canine rabies

Participants in both study areas correctly described the most widely-recognized symptoms of canine rabies: dog aggressiveness, excessive drooling or foam from the dog’s mouth, and a change in the dog’s behavior. Some participants could describe less well-known symptoms: bright, red, wide-open eyes; staggering; disorientation; and hydrophobia or unwillingness to drink water. No participants recounted the paralytic or dumb form of rabies (in which rabid dogs exhibit progressive limb weakness and inability to swallow, with people frequently presuming dogs have something stuck in their mouths). Participants’ knowledge about what a rabid dog looked like seemed to affect their treatment seeking decision-making for a dog bite if the dog seemed well:

> *“Because I see there’s nothing wrong with my neighbor’s dog, so it is not worth going (to get treated) and because it takes time. What for, if the dog is fine? [If the dog shows rabies signs later], then yes*.*” (Man, peri-urban area, owner)*

Participants could correctly describe that, following infection with the rabies virus, one might not have any symptoms for days or weeks. While almost all participants thought incorrectly that canine rabies can be easily diagnosed with a simple blood test, only two—from the urban area—mentioned that rabies attacks the dog’s nervous system and that it is necessary to analyze the dog’s brain to confirm disease. Urban residents indicated that they might only seek treatment if signs of infection or other symptoms possibly related to a dog bite or to rabies emerged— focusing on bites with significant soft-tissue damage, but not worrying about small skin-breaking bites.

### 3.3. Knowledge of and barriers to management of dog bite wounds

FGD participants recognized dog bites as a mode of transmission of rabies virus to humans, and could identify the three key preventive steps that are posted in most area health facilities (Figure 2): 1) wash the wound with water and soap; 2) identify the dog; and 3) seek medical care and report the offending dog at the nearest health post/health center. Participants identified health communication material (Figure 3) as the source of this knowledge.

**Figure 1.**
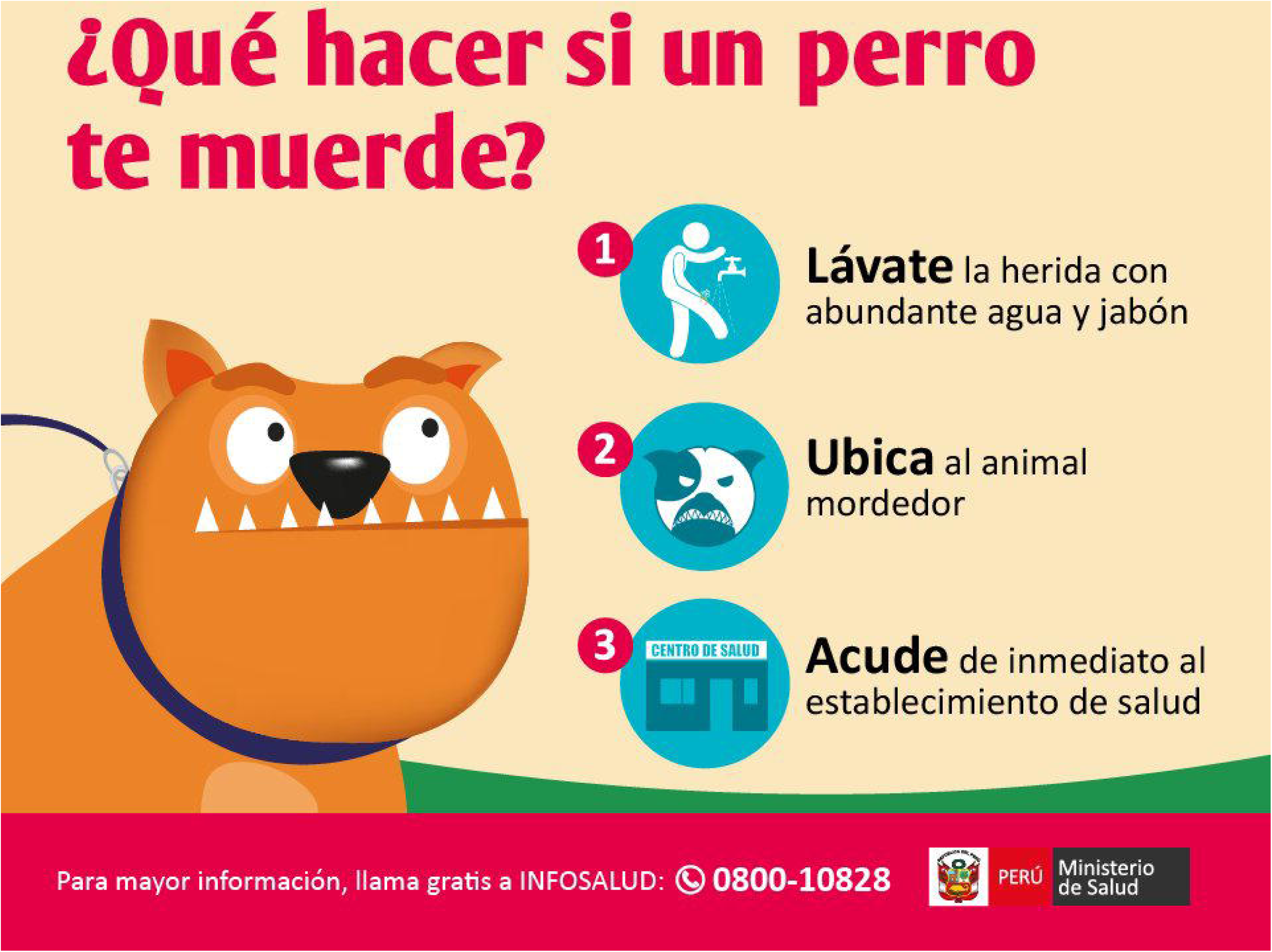
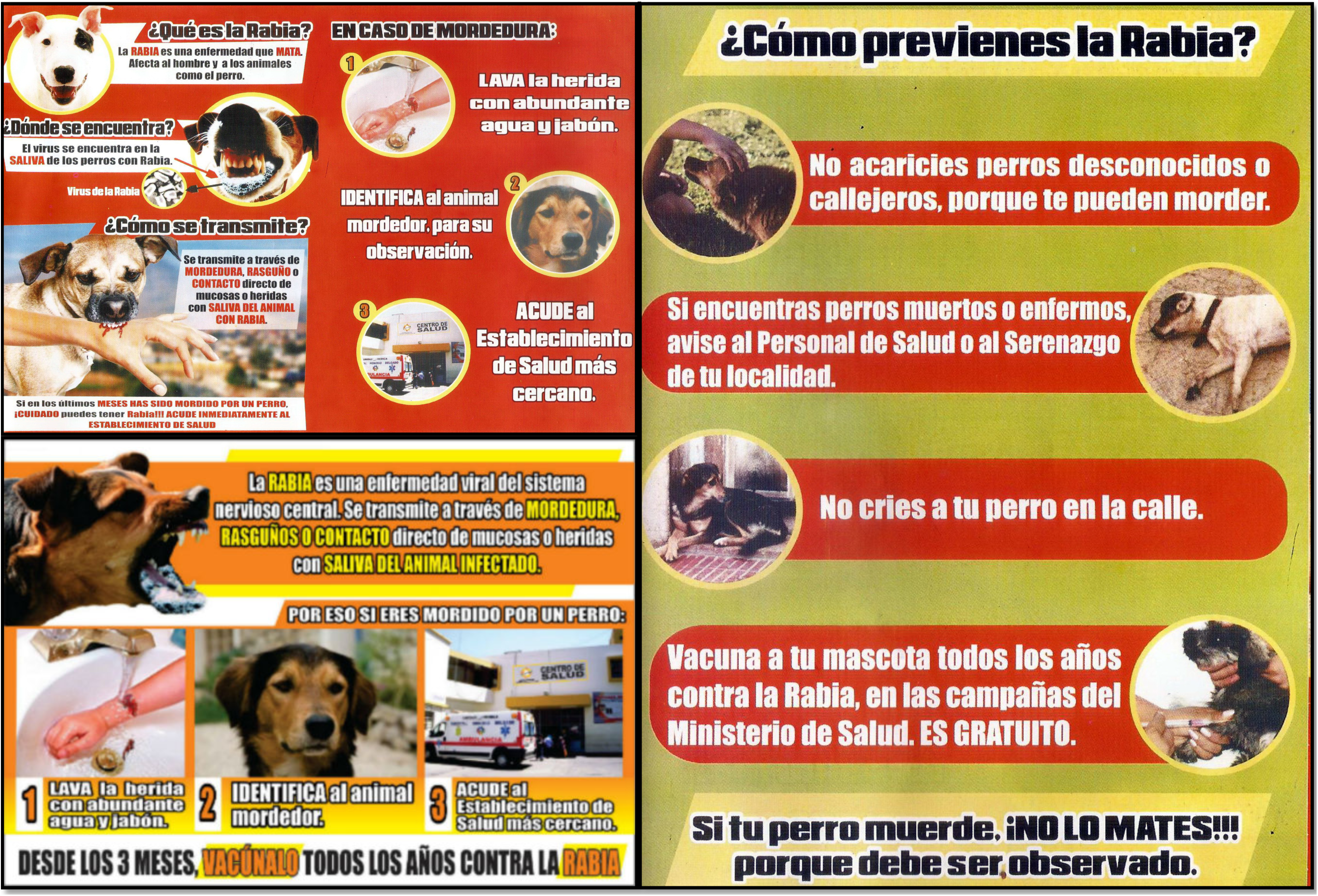
(New photos of urban and peri-urban areas)

Despite knowing what to do, most respondents reported not taking action themselves or knowing people who had not sought care or reported a dog following a bite. The most common stated reasons for not seeking care were: 1) lack of awareness that a small bite or skin tear require care, 2) poor treatment at the health post or health center, 3) logistical and economic barriers to seeking care, 4) use of alternative treatments, and 5) perception that a “known” or “good” dog is “safe”.

Respondents agreed that few people would seek care for a minor scratch or bite unless there was significant pain, bruising, bleeding, broken tissue, or the wound failed to heal on its own:

> *“When the wound bleeds too much, when it doesn’t heal and when it’s way too serious - that’s when you go. When it’s a small bite, people leave it; they cure it with alcohol and leave it*.*” (Woman, peri-urban area)*

Respondents did not draw a connection from waiting to see if the wound healed on its own and delayed treatment.

Perceptions about and experiences of poor quality care was another barrier to seeking care following a bite. Most respondents live near a health post (health centers and hospitals are further away), but had more negative opinions about the posts, particularly the limited hours these are open, the poor customer service, and the stock-outs of the rabies vaccine (which our team confirmed at a nearby health facility following the FGD):

> *“They mistreat us, don’t attend to us… [Those doctors say] ‘oh, there’s no pills here’. From the post they send us down to the health center, and there they ask us ‘Why were you sent here? They have doctors up there, the health post has this and that…’” (Woman, peri-urban area)*

Logistical and economic reasons (i.e., distance to health facility, lack of time, and perceived cost) also prevented respondents from seeking care in a health facility after a dog bite, primarily in the peri-urban area. Participants noted that going to the health post involves hours or even a whole working day, which was not considered feasible, particularly when a dog bite is considered treatable at home:

> *“It’s a waste of time going to the health post when we can treat it, based on our experience, with soap, hydrogen peroxide…. and it’s cured!” (Woman, urban area)*.

Some male participants explained that they could not waste work time seeking medical care in the health posts (this comment from a peri-urban male elicited several nods from other men in the group):

> *“I wouldn’t go because I have debts, I have to work and the post is often full so there’s no time, you waste time—one, two, three hours—hours that you lose of work, so I prefer not to go. You don’t really get a sense of being at risk or not*.*”*

Participants revealed a preference for alternative treatments for dog bites that also reduced likelihood of going to the health post for care. In urban areas people mentioned other remedies such as soap, hydrogen peroxide, penicillin or Mentholatum. People in the peri-urban areas were more likely to mention alternatives associated with local popular healing practices, such as aloe vera and herbs, and even burnt dog hair:

> *Woman: When I was little a dog bit me on the leg and made a deep wound. The owner cut the dog’s hair, burnt it and put it on the sore with a patch. It scarred well*.

> *Man: That’s a grandmother’s secret. (Peri-urban area)*

Older adults were reported as being more likely to use local healing practices—ranging from Mentholatum to herbs to hair ashes—and more reticent to seek care at a health facility than younger people.

Finally, there was also the perception—in both urban and peri-urban areas—that seeking care for a dog bite depended on the specific dog. When probed during the FGDs, peri-urban residents were more likely than urban residents to report having ever been bitten and having been bitten in the past year (some reported three or four bites in the past year); they were also more likely to report not seeking care following a bite. If the biting dog was their own animal or if the owner was known to care for the dog, participants who had been bitten were less likely to seek care. If they did not know the dog’s owner, they perceived a higher chance of acquiring diseases, including rabies. The dog’s behavior also influenced their assessment of rabies risk and subsequent response to a dog bite. If the biting dog was “fierce” (“*perro bravo*”) or showed aggressive behavior, even if the wound was small, and the dog had an owner, they were more likely to go to the health post than if it was a less aggressive dog. A woman in the peri-urban FGD mentioned that her once-gentle dog suddenly became aggressive, but she did not relate the behavior change to rabies, nor the need to seek treatment for the scratches:

> *Woman: … and (the dog) started biting my husband. One day she was in my room acting crazy, started to bite and bark (*…*) I was watching TV, I hadn’t done anything to her, she was irritated (…) and she started biting at me “woof woof woof”. She bit me and I hit her head with my TV thinking she would run away but she didn’t and kept biting me. [It occurred] a month ago I think. My husband had to kill her because she got like that*.

> *Facilitator: Where did she bite you?*

> *Woman: On the leg, but it was just a scratch, she didn’t do anything to me*.

> *Facilitator: Was there any blood?*

> *Woman: Yes (…)*

> *Facilitator: Did you go to the health post?*

> *Woman: No, I washed it with boiled water and soap*.*\**

### 3.4. Knowledge about and practices associated with post-exposure prophylaxis (PEP) and the importance of its completion

When asked about barriers to accepting PEP or completing PEP once started, participants identified six reasons: 1) rabies vaccines are too painful, 2) people thinking that multiple vaccines could be harmful, 3) people feeling that one or two vaccines are enough, 4) people not wanting to complete the series of vaccines because they are not treated well at health facilities or by health personnel, 5) people not having appropriate information about the importance of completing PEP, and 6) lack of time to continue with the PEP regime. Participants from the urban areas emphasized the first, second, and third responses, while those from peri-urban areas were more likely to mention the fourth, fifth, and sixth reasons.

Participants reported that the rabies vaccine has a particular reputation for being painful. FGD participants in the urban area specifically (and a few in the peri-urban area) conjectured that people might avoid the rabies vaccine for that reason. Poor skills in administering the vaccine was raised as a cause of pain. Estimates of the number of doses of the vaccine required for prophylaxis varied widely, with some older participants mentioning 20 or more shots (as was the case in the 1970s).

Belief that the full series (5 shots) could be harmful was cited as a reason to not adhere the complete series. Multiple respondents noted that “the whole dosage” could have adverse effects, and that other people say that receiving too many vaccines is dangerous:

> *“I have heard that if you get all the vaccine doses, it harms people…That is why they don’t want to be given the vaccines; other people tell them not to [get the full series] because it’s harmful*.*” (Woman, urban area)*.

Participants reported others might stop PEP early if there is no evidence of symptoms and the wound is healed (from start to end of vaccination, it would be 4 weeks). They compared completing PEP to completing antibiotics when one is ill: many stop taking antibiotics when they feel better. One woman in the peri-urban area got all her vaccines when she was bitten, but admitted that if her leg had not looked so swollen, she would have stopped going after the first few vaccines:

> *“I suppose the person tells the doctor ‘But why should I have five vaccines if I’ve already had three? Besides, I don’t feel anything, it doesn’t hurt, doesn’t itch, doesn’t sting, nothing, I don’t have anything, or don’t have any discomfort’*.*”*

Only peri-urban participants mentioned the quality of services at health facilities as a barrier to continuing PEP. Perceptions of mistreatment surfaced numerous times:

> *Woman1: …what am I going to go for? To be treated like that? I’d better not go, I’ll just take care of myself*.

> *Woman2: The same with me, they told me ‘no, no, no, you go somewhere else to get attended’. (Peri-urban area)*

Suspicions about vaccine quality were also raised, but respondents could not offer further details.

In both areas, participants perceived a lack of information about the risks of discontinuing the PEP regimen, which in turn reduces motivation to make time for dog-bite related medical treatment; participants also referenced people’s ignorance or carelessness in dealing with important issues.

> *Man: [They don’t continue their vaccines] because people work, they have their families. Facilitator: Even though they can die?*

> *Woman: There are very ignorant people who don’t even care what happens to their children and the rest of people, they just don’t care. (Peri-urban area)*

In both areas, lack of time was cited as a likely reason for not seeking to complete one’s vaccination series, specifically referring to people not wanting or not being able to take time off of work. In some parts of Arequipa, this argument might be unfounded since health workers actually go to the homes of people receiving PEP vaccination. Some women in the peri-urban area also mentioned that women might not go due to child care needs. Moreover, because the health posts that primarily service peri-urban communities may not offer vaccines, time spent seeking care at a health center is generally higher for those in peri-urban areas and participants complained about this.

## 4. Discussion

An epidemic of canine rabies in Arequipa has received widespread attention, from radio and TV messages to posters at health facilities (Figures 2 and 3) and it is clear that some specific knowledge gaps persist. For example, most FGD participants correctly associated aggressive and salivating dogs with rabies, but none identified the paralytic form of the disease, which is similar to what others have found (12,13,26,32,33). It is important to increase awareness of this common (34) clinical manifestation of rabies that may increase human exposure to the virus but may be missed by the community and the health system. Most participants also knew that dogs could get rabies from other dog bites; however, many others thought dogs could get rabies from being kept on rooftops or other hot places, unattended, with insufficient water, similar to what has been reported in Ethiopia (35). Some thought that flies can transmit rabies, similar to what was observed in Bali (36). Other reported misbeliefs such as the inhalation route (35) or any contact with dog saliva regardless of skin integrity (27,35), were not stated by our participants. It is very concerning that two participants in our study experienced a clearly risky scenario (i.e., dog behavior change, aggression towards members of the family, a bleeding bite), but killed and buried the dog and did not seek medical care or notify authorities. Other participants mentioned disposing of dead dogs in the same way without reporting them to health inspectors, suggesting lack of community awareness and engagement with the rabies surveillance system.

While these knowledge gaps may be partially responsible for poor prevention-related behaviors, there were additional behavioral drivers of failure to adhere to prevention guidelines among those who who could articulate the importance of seeking care and other recommended actions post-dog bite. Several participants displayed common mental shortcuts, cognitive biases, or other psychological factors that made it difficult to act on intentions to follow prevention guidance following a potential exposure. For example, some participants downplayed the risk of their own dog contracting rabies, even when acting atypically aggressive. This is likely due to some combination of optimism bias (37,38) myside bias (39), and halo effect (40), all of which will contribute to misperceptions about the relative risk of one’s own dog – a familiar animal for whom on may have fond feelings – contracting rabies vs. a stray dog, and the assessment of one’s own risk for contracting rabies from a familiar vs. a strange dog.

Other behavioral barriers to appropriate prevention are related to mental models (41) about rabies and prevention, some of which may be inadvertently supported by preventive promotional material and some of which come from long-standing folk wisdom. For example, the perception that one need not seek care for a superficial bite or scratch, or a bite from a seemingly-healthy dog, may have been influenced by posters at health facilities that show extremely aggressive-looking dogs foaming at the mouth, and images of severe, bloody bites on an arm. While the graphic quality of these images is attention-grabing, it may have had the unintended consequence of anchoring people’s perceptions of what a rabid dog is like and the severity of bites that require attention. Another very common bias—present biased-preferences or hyperbolic discounting (42) can explain how even minimal hassle factors drive nonadherence to PEP once recommended (43). Because we overweight immediate costs and benefits of a behavior, and heavily discount consequences that are probabilistic or in the future, the hassle and inconveneience of repeated trips to a crowded clinic can easily outweigh the perceived future health benefit of completing the series.

A third set of barriers to effective secondary prevention identified in our study is related to trust in and perceived quality of health services. Many participants had lost faith in the ability of their local health posts to attend to them. During an emergency as this one, all health facilities – *particularly* the small ones in the more remote peri-urban areas – must be equipped and prepared to initiate and provide PEP to individuals who require it. Referring exposured individuals away means they may be delayed, or never, receive PEP. As an example, when the research team checked with healthcare providers at a nearby health care facility equipped with a refrigerator, in an area with several cases of verified canine rabies, they were told that, as a small facility, the health post did not have any human rabies vaccines and people should seek care at a larger health center that was a difficult (due to hilly terrain) 30-minute walk away.

Completing the course of PEP once prescribed also emerged as a gap in effective rabies prevention. Our health authority collaborators explained that they do reach out to individuals who have not completed a vaccination series, but sometimes individuals refuse to see them or pretend they are away. While this might be due to a knowledge gap about disease severity, or hassle factors, there may also be other behavioral biases at play: Direct experiences or anecdotes of previous mistreatment at health facilities may contribute to reactive devaluation (devaluing the ideas or proposals that come from an antagonist (44) of health professionals’ recommendaitons. Information avoidance or “ostriching” (45) may also lead people to purposefully avoid or tune out upsetting information such as rabies disease risk or the demands of effective prevention.

Our results point to solutions that could close the rabies prevention gap for human populations leaving in canine rabies endemic areas. Using media appropriately is a proven strategy to increase health service utilization [40], and finding the best communication channels will allow targeting more effectively specific populations. Moreover, to improve knowledge, messages must be appropriate for the audience they are intended for, and piloted and tested to ensure comprehension. Health education messages need to be clear that any type of dog can get rabies—whether it is a stray dog or a loved and well-cared-for house dog—all it takes is one small wound. While seeking care for a “mere” scratch might feel silly, or like a waste of time, it is critical in the midst of an outbreak that public health messaging around rabies motivate individuals to seek care even if the bite seems superficial or trivial.

However, some of the problems identified appear to be driven by biases underlying both intentions and behaviors that are unrelated to knowledge. For example, most individuals have experienced a dog bite at some point and known others experiencing dog bites where “nothing happened” despite being bitten—and disregard (or choose to disregard) that the circumstances are different when there is a canine rabies epidemic in the city. Behavior change communication campaigns and program improvement efforts alike can incorporate awareness of these cognitive biases and design both messaging and health services to counter them. For example, default stocking of rabies vaccine at all health posts and “fast track” treatment for patients arriving following an exposure would reduce real and perceived hassle factors.

There are some important limitations to this qualitative study. We used purposive sampling and our participants were recruited during daytime hours; hence, it is not a comprehensive report of the views of others in the community. However, by the 7th and 8th FGDs, there was a saturation of themes that emerged for the various topics of interest. Although our team took note of health promotion materials that were visible in the health facilities or community, our study did not examine all sources of health education (or possible sources of misconceptions) in Arequipa. Questions regarding possible reasons for lack of PEP vaccination completion were asked of community members, based on what they have heard and observed in their communities regarding the rabies vaccine, but not the patients who had not completed their PEP vaccination regime; including this group would have been ideal, but outside the scope of the design of this study.

## 5. Conclusions

Our study describes knowledge deficits and behavioral biases affecting preventive practices for rabies in the city of Arequipa, Peru. We identify health education topics that should be emphasized to improve rabies prevention, common misperceptions to be addressed by communication campaigns, as well as suggest possible behavioral biases that could be addressed programmatically. To close knowledge gaps, salient messaging is urgently needed on the following topics: rabies is fatal for humans and dogs and not treatable once the clinical signs and symptoms start; any dog bite, even a small one from a known and seemingly healthy dog, can transmit the rabies virus, therefore seeking medical care is necessary to assess one’s risk after a dog bite; if PEP vaccination is indicated, it is because the person is at risk of rabies; and, rabies fatality can be prevented if PEP vaccination is started promptly and is completed. Health practitioners could play an important role in promoting trust in their patients and communicating how the benefits of PEP vaccination outweigh the small cost of trips to the health facility and tolerable pain caused by injections; they could also reduce the steps and hassle for patients to receive treatment; for example, health posts in peri-urban locations must stop referring patients to other health facilities when these health posts could be equipped to vaccinate and manage patients within their localities to increase likelihood of patients seeking and obtaining care. Analysis of the gaps in knowledge as well as the ingrained cognitive biases surrounding canine rabies and health posts in the Arequipa community suggests that targeted strategies by level of community urbanization may be beneficial in dissolving barriers to PEP during a rabies epidemic.

## Data Availability

The data that support the findings of this study are openly available.

https://github.com/chirimacha/Rabies_PEP_qualitative_spatial_inequalities

## Acknowledgments

This work was supported by the Eunice Kennedy Shriver National Institute of Child Health and Human Development (grant number R01HD075869). We acknowledge the work of the members of the One Health Unit for their contribution to the implementation of this study, especially Lina Mollesaca and Carlos Condori.

## Author Contributions

R.C.N., M.Z.L., A.M.B., and V.A.P.S. conceived and designed the experiments; J.Br., R.C.N., V.A.P.S., and K.B.M. conducted focus groups; J.Br., C.A.N., K.B.M, and V.A.P.S. analyzed the data; M.Z.L., J.Br., and A.M.B. administered and supervised the study, V.B. contributed with resources including facilities; and R.C.N., J.Br., A.M.B., and V.A.P.S. wrote and reviewed the paper.

